# Effectiveness of Switching CGRP monoclonal antibodies in non-responder patients in the UAE: A retrospective study

**DOI:** 10.1101/2023.08.04.23293651

**Authors:** Reem Suliman, Vanessa Santos, Ibrahim Al Qaissi, Batool Aldaher, Ahmed Al Fardan, Hajir Al Barrawy, Yazan Bader, Jonna Lyn Supena, Kathrina Alejandro, Taoufik Alsaadi

## Abstract

Calcitonin gene-related peptide monoclonal antibodies (CGRP mAbs) have shown promising effectiveness in migraine management compared to other preventative treatment options. Currently there are several studies related to the efficacy and tolerability of CGRP mAbs in the management of mgraine. However, many questions remain unanswered when it comes to switching between antibody classes as a treatment option in patients with migraine headaches. The present study seeks to explore and assess the treatment response to CGRP mAb in patients who have previously failed other CGRP mAbs.

This was a retrospective, real-world, exploratory study. The participants included within the study were adult (≥18 years) patients diagnosed with migraine. Patients who were treated with two or more GCRP mAbs were retrospectively analyzed. Data was collected from one site, 53 patients with migraine headache switched between three CGRP mAb types (Eptinezumab, Erenumab, and Glacanezumb) due to lack of efficacy of the original prescribed CGRP mAb. Efficacy of switching between types of CGRP mAb’s was evaluated through documented MMD’s in patient diaries and clinical records. Non-parametric analysis was used to compare efficacy of the first six months of each prescribed medication. The analysis of efficacy demonstrated that some improvements were seen in both class switch cohorts (CGRP/R to CGRP/L and CGRP/L to CGRP/R). However, the most noticeable improvement in efficacy of the prescription switch was found in patients who switched between different medications of the CGRP/L class. Both chronic migraine and episodic migraine patients showed improved MMD’s, however chronic migraine patients demonstrated higher responsiveness of efficacy following this lateral switching, The safety of switching between CGRP classes was well observed as any adverse events presented pre-class switch did not lead to the discontinuation of treatment following the later switch. The findings of this study suggest that switching between different classes of CGRP mAbs is a potentially safe and clinically viable practice that may have some applications for those experiencing side effects on their current CGRP mAb or have suboptimal response. This is especially true for patients initiating treatment on ligand targeted CGRP mAb who experience side effects or lack of meaningful efficacy, as the ligand-ligand cohort seems to demonstrate the best outcome. Larger cohort studies and longer follow ups are needed to validate our findings.

## Introduction

Migraine is a neurological disorder experienced by an estimated global point prevalence range of 14,107 cases per 100,000 (1). According to the 2019 Global Burden of Disease Study, migraine is the second most common non-fatal disease in terms of “years lived with disability” (2). Those suffering from migraine experience a significant impact on their ability to maintain their productivity and relationships, mandating continuous efforts to better understand its pathophysiology and optimal treatment options (3). Although the precise mechanism of migraine remains unknown, recent findings suggest the calcitonin gene-related peptide (CGRP) plays an integral role (4). Responsible for nociception within the trigeminal ganglion, CGRP represents a major point of intrigue in the development of migraine prophylactic medication. Thus, numerous studies have emerged within the past decade investigating the effectiveness of monoclonal antibodies (mAbs) as CGRP receptor antagonists in migraine treatment.

Older treatment options for migraine prevention have not been very successful in alleviating the personal and economic burden of migraine (5). A major reason for the lack of success is their limited tolerability and patient adherence (6). Antiepileptics, beta-blockers and antidepressants are examples of these medications. Furthermore, these medications have not been very effective in treating migraine headaches and reducing migraine burden (6).

On the other hand, mAbs result in better treatment outcomes and, due to their long half-lives, they can be dosed at long intervals, which can be a preferable option for some patients (7). Less frequent dosing can minimize the burden on the patient and assures better treatment adherence. Presently, a small collection of monoclonal antibody medications have been approved for the preventative treatment of episodic and chronic migraine by the U.S. Food and Drug Administration (FDA). Such monoclonal antibodies include: Galcanezumab, Eptinezumab and Fremanezumab which target the CGRP ligand and Erenumab which targets CGRP receptors Table 1 (4).

**Table 1:**
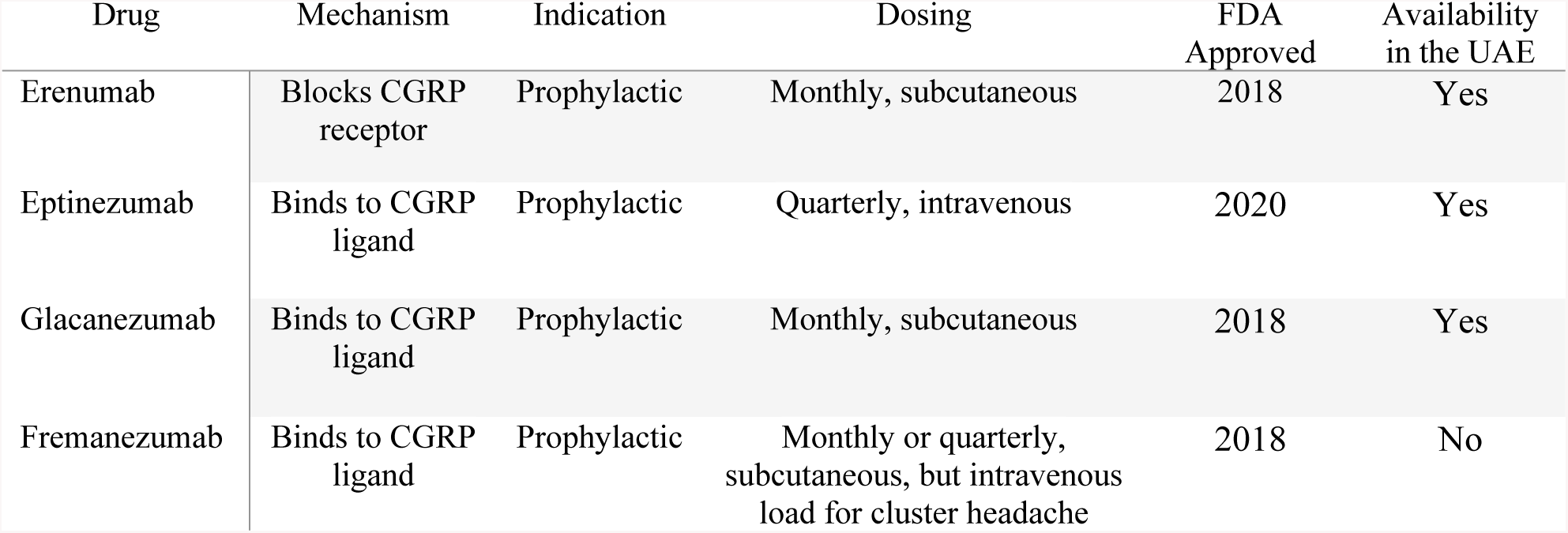
CGRP Targeting drugs.

Recent studies examining usage of Erenumab suggest that it can effectively reduce migraine frequency and improve quality of life (8). In a similar study assessing the efficacy and tolerability of Erenumab among 418 patients, 168 (69.7 %) reported that the benefits of Erenumab outweighed any potential drawbacks (9). Episodic migraine patients with at least one previous preventative treatment failure (PPTF) exhibited significantly greater gains in efficacy compared to placebo (10). Among the aforementioned mAbs, Eptinezumab stands out as the sole drug capable of intravenous administration, allowing for a rapid onset of action (11). This unique quality allows for quicker headache pain relief (12), and reduced monthly migraine days compared to placebo (13).Similarly, the effectiveness of Galcanezumab was well established for patients with episodic and chronic migraines (14). Existing literature on CGRP monoclonal antibodies (CGRP mAbs) suggests high patient tolerability; discontinuation is most commonly attributed to lack of efficacy rather than adverse effects (15). Despite a lack of major differences in efficacy across clinical trials, a few case series studies found some patients who have suboptimal response to one mAb has managed to successfully switch to another one with noted significant improvements (16–19). One real-world analysis study demonstrated that one-third of Erenumab non-responders achieved >30% response after switching to another CGRP-mAb (20).

These findings warrant further investigation into the efficacy and safety of switching between CGRP-mAb classes. Therefore, this study will investigate the treatment effectiveness, tolerability, and adherence of migraine patients in the United Arab Emirates (UAE) following a switch from a PPTF to another mAb. This study will serve as an integral step toward advancing our understanding of migraine treatment. It shall also represent the first of its kind by focusing on an underrepresented population in clinical research.

## Methods

### Study Design

This was a retrospective, real-world exploratory study. Data used was gathered from one site, The American Center for Psychiatry and Neurology (ACPN), Abu Dhabi, UAE. A total of 391 patients with episodic migraine (EM) or chronic migraine (CM) who have received at least one dose of GCGRP/R mAb (Erenumab), or CGRP/L mAb (Eptinezumab, or Glacanezumab) were reviewed for eligibility to be included in the study. Fremanezumab is currently not available in the UAE, therefore it has not been included in the analysis.

Data was gathered from patient’s clinical records which contain all the required demographic information, diagnosis, medication history, Monthly Migraine Day (MMD) at baseline and at follow up visits. Additionally, patient satisfaction with medication was documented in their clinical records. Patients were asked to keep a record of attacks and symptoms in their headache diaries. Efficacy of the prescribed treatment was evaluated by measuring the change in MMD between visits. Safety was also assessed; this was done by monitoring adverse events.

Follow-up visits were scheduled monthly or as deemed necessary, which is a standard protocol at our site for all patients initiating treatment on mAbs. Patients were assessed on their baseline frequency of MMDs and, subsequently, thorough discussions with treating physician on which mAb would be most effective, addressing each patients’ specific needs. If the current medication did not result in any meaningful reduction of MMDs, an option to switch to another mAb was offered to the patient.

The retrospective analysis mainly focused on two main periods. The first period included data of patients treated with a specific CGRP mAb, while the second period involved data on patients who switched to another anti CGRP mAb. Switching was mainly due to lack of efficacy; it was ensured that patients included in the analysis completed at least 3 months of treatment before switching. During each phase, patients’ MMDs were assessed at 3 stages prior to and following their medication switch: at least one month before the first injection (baseline), at a 3-month follow-up, and at a 6-month follow-up.

### Ethics

This study was conducted in accordance with the Helsinki Declaration of 1964 and consistent with Good Clinical Practice (GCP). All ethical guidelines, health authority regulations and data privacy laws were ensured. Prior to the start of the study all relevant approvals were obtained from ACPNs Institutional Review Board (IRB), a waiver of informed consent from the corresponding ethics committee was obtained. To ensure transparency and accuracy all authors were given access to the study data.

### Sample

Records from ACPNs nursing department were gathered where all patients who have been administered one mAb have been screened and identified (Figure 1). Data included patients from January 2018 up to September 2022, who are adults (≥18 years), and who had a diagnosis of either EM or CM, as per the International Classification of Headache Disorders (ICHD-3) criteria. Data was shared independent of treatment effects or cause for the GGRP mAb switch.

**Figure 1:**
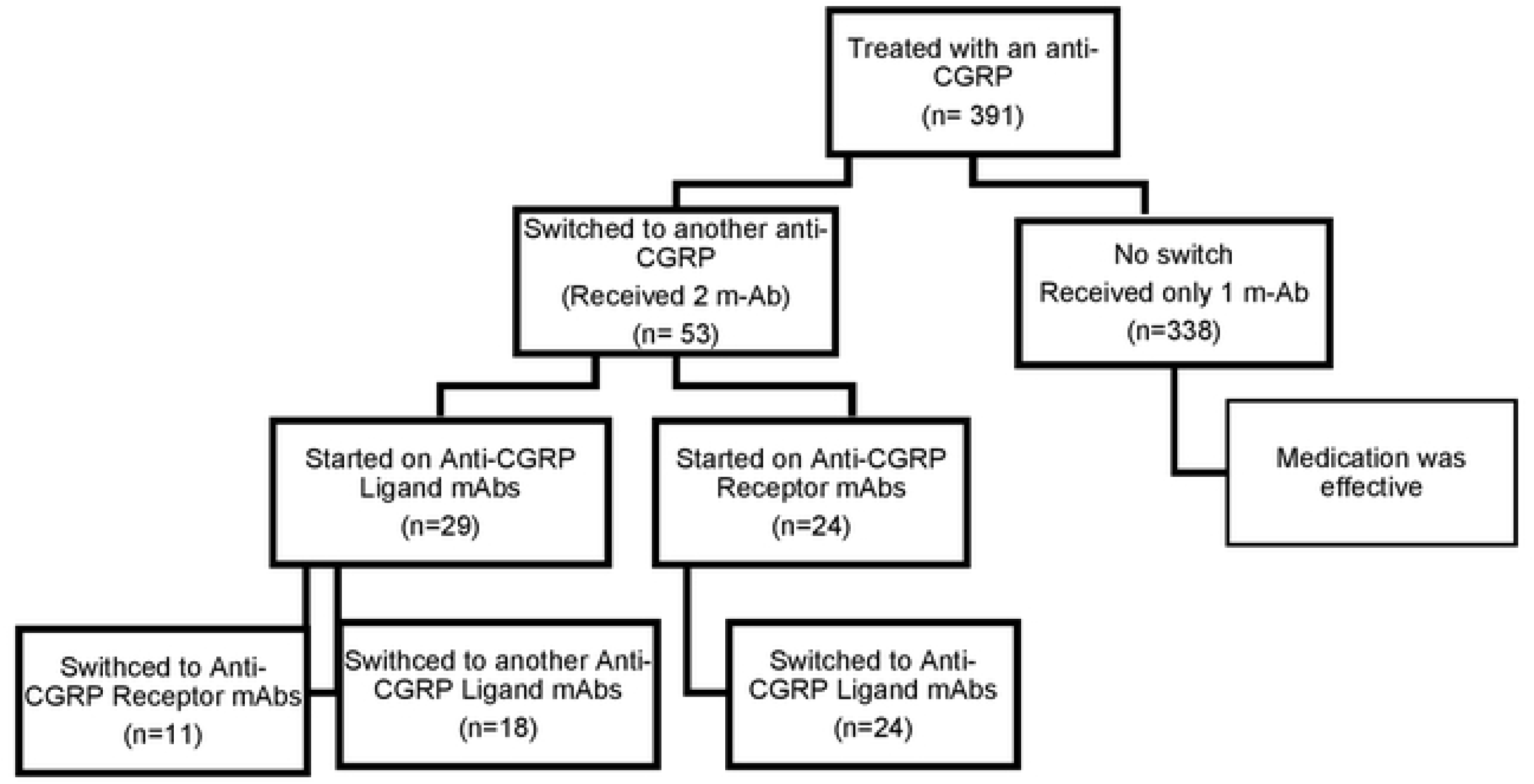
Flowchart of patients.

Patients were included in the analysis if they (i) switched between two of the previously mentioned CGRP mAbs (switchers) (ii) received at least three doses of the first CGRP mAb and maintained treatment for a minimum of 6 month after switching, Those who have demonstrated a meaningful response, which is defined as more than 50% reduction in MMD for EM, and more than 30% for CM, and were satisfied with their treatment remained on their current preventative treatment hence they were not included in the effectiveness analysis. Patients were categorized according to their switching profiles. The three profiles represented were: CGRP/R mAb to GCRP/L mAb, CGRP/L to CGRP/R, or CGRP/L to another CGRP/L mAb (Fig 1).

### Variables, data extraction, and endpoints

The number of MMDS were extracted from the headache diaries as documented on the patients’ electronic medical record (EMR). Due to the non-standardized headache diaries and the varying details of documentation of headache characteristics and accompanying symptoms during each headache attack, reliable differentiation between headache and migraine days was not possible. The primary endpoint was the absolute change from baseline in MMDs response rate (>25%, >50%, >75%, and 100% reduction in MMDs) of each category of the switchers. As per the methodology outlined by Kaltseis et al. in 2023, patients who demonstrated a positive response to treatment were classified as responders. This was determined by a minimum reduction of 50% in MMDs for EM or a minimum reduction of 30% in MMDs for CM after receiving treatment for a minimum of 3 months. Patient characteristics including age, gender, migraine diagnoses, migraine years, and type of CGRP-mAb from the EMR.

### Objectives

The primary objective was to retrospectively assess the reduction in frequency of migraines and determine the efficacy of the current preventative treatment following the switch from a previous CGRP mAb. Additionally, the study sought to evaluate the tolerability of the treatment and report any adverse reactions.

### Statistical Analysis

Since this analysis was conducted retrospectively the sample size was not based on any statistical consideration. The sample size was achieved depending on the number of cases fulfilling the inclusion criteria treated at ACPN. Continuous variables were summarized using mean ± standard deviation [SD] or median interquartile range [IQR], while categorical data were presented as numbers and percentages. The normality assumption was evaluated using the Shapiro-Wilk test. Given that the data did not follow a normal distribution, the Wilcoxon signed-rank test was used to analyze the changes in quantitative variables pre-post changes. A significance level of p < 0.05 was considered statistically significant for all variables. The statistical software SPSS version 26.0 (IBM Corp. SPSS Statistics, Armonk, NY, USA) was utilized for all data analyses.

## Results

### Demographic and Baseline Characteristics

The participant pool was composed of 53 individuals; all of whom had undergone a switch from one CGRP mAb to another; the descriptive statistics for this cohort are visualized in **Table 2**. Among the 53 participants, 42 (79.2%) of whom were female, 20 (37.7%) were diagnosed with CM while the remaining 33 (62.3%) were diagnosed with EM. The mean age (SD) of participants in years was 39.2 (11.0). Furthermore, patients were categorized according to their switching profile, they had the following distribution CGRP/L to R mAb (n=11; 20.7%), CGRP/R to L mAb (n=24; 45.3%), and CGRP/L to L mAb (n=18; 34.0%).

**Table 2:**
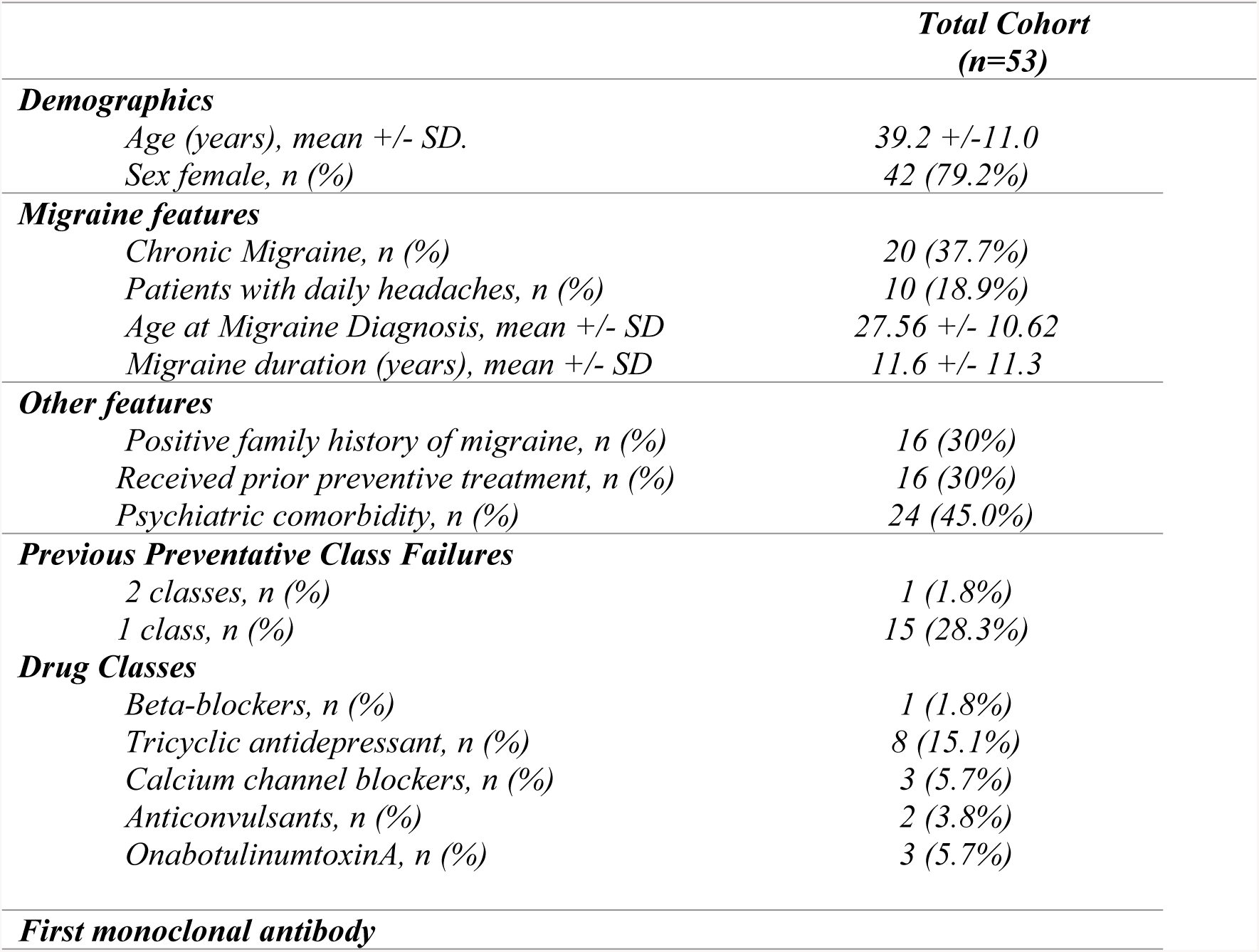
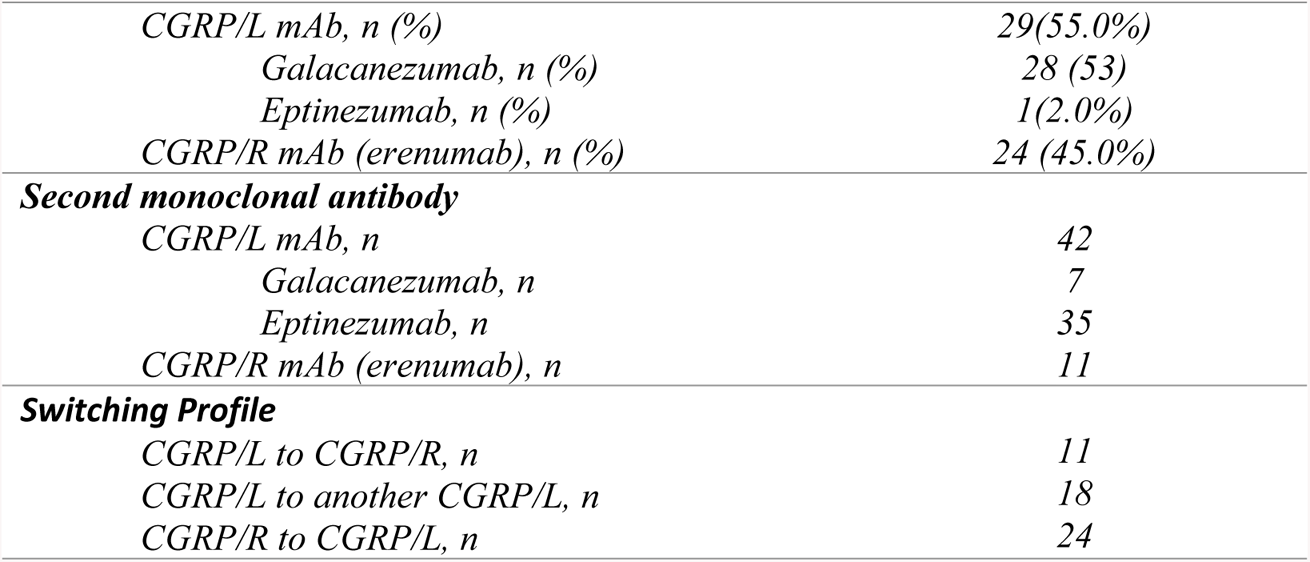
Patient demographics and clinical features.

Out of the 53 patients, 16 individuals had previously attempted preventative treatment for migraine. One patient has tried two preventative therapies before switching to a CGRP mAb, while the remaining 15 only had one preventative treatment failure prior to switching. Previous preventative treatments included: Propranolol, Amitriptyline, Flunarizine, Topiramate, and OnabotulinumtoxinA **Table 2**.

Throughout the study, the frequency of participants’ monthly migraine days (MMD) was assessed at 3 stages prior and following their medication switch: at baseline, at a 3-month follow-up, and a 6-month follow-up, this can be visualized in **Fig 2**. As exhibited in **Table 3**, the greatest mean (SD) MMD values were recorded at the baseline assessments; however, it is worth mentioning that the post-switch baseline mean of 10.21 (5.42) was lower than that of the pre-switch baseline mean of 12.92 (8.23). Following the pre-switch baseline assessment, the mean was reduced to 5.53 (5.73) by month 3, however it rose slightly to 5.92 (6.69) by month 6. Following the post-switch baseline assessment, mean MMD dropped to 5.74 (4.32) by month 3 further decreased to 5.42 (4.98) by month 6.

**Figure 2:**
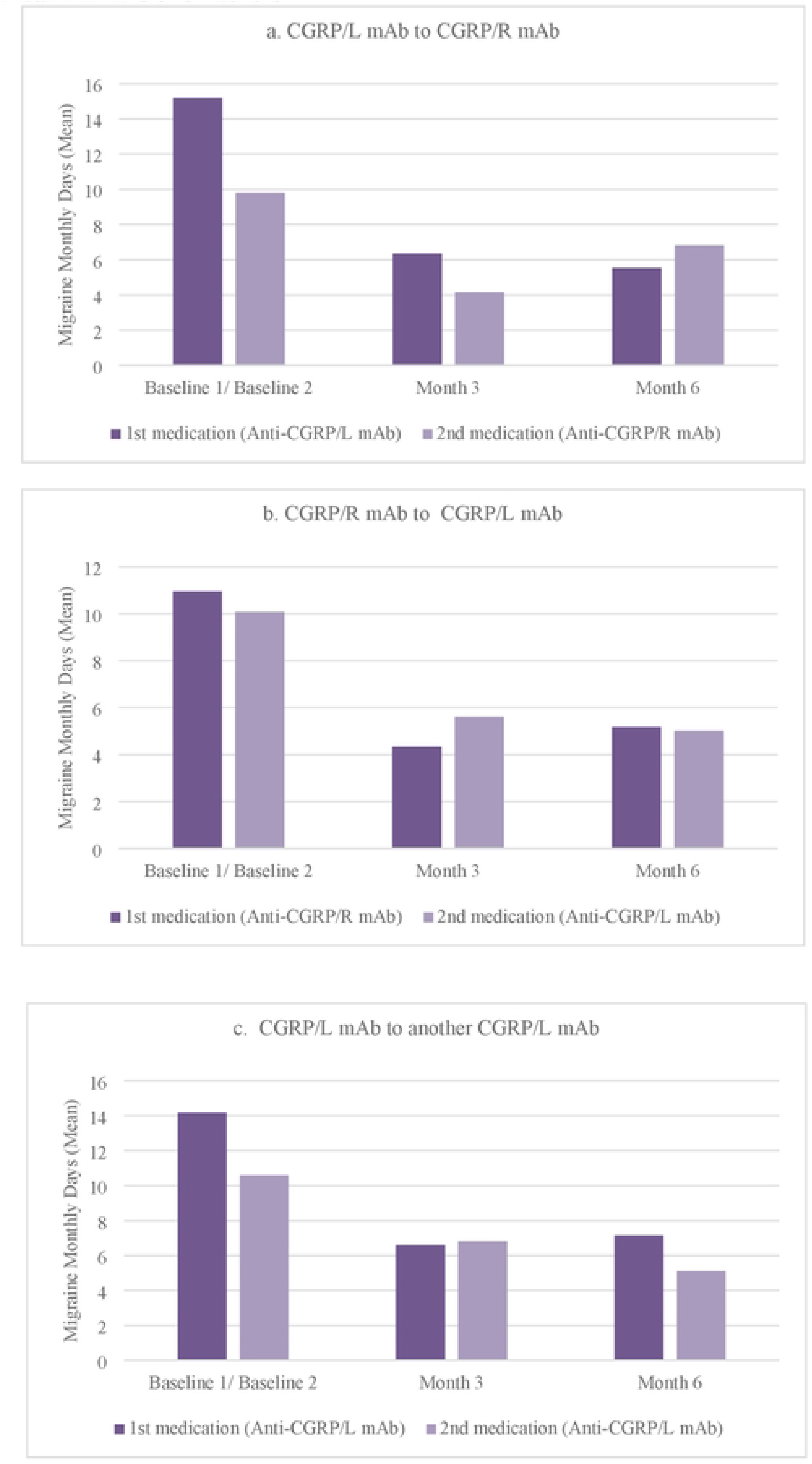
Mean MMD’s of switchers.

**Table 3:**
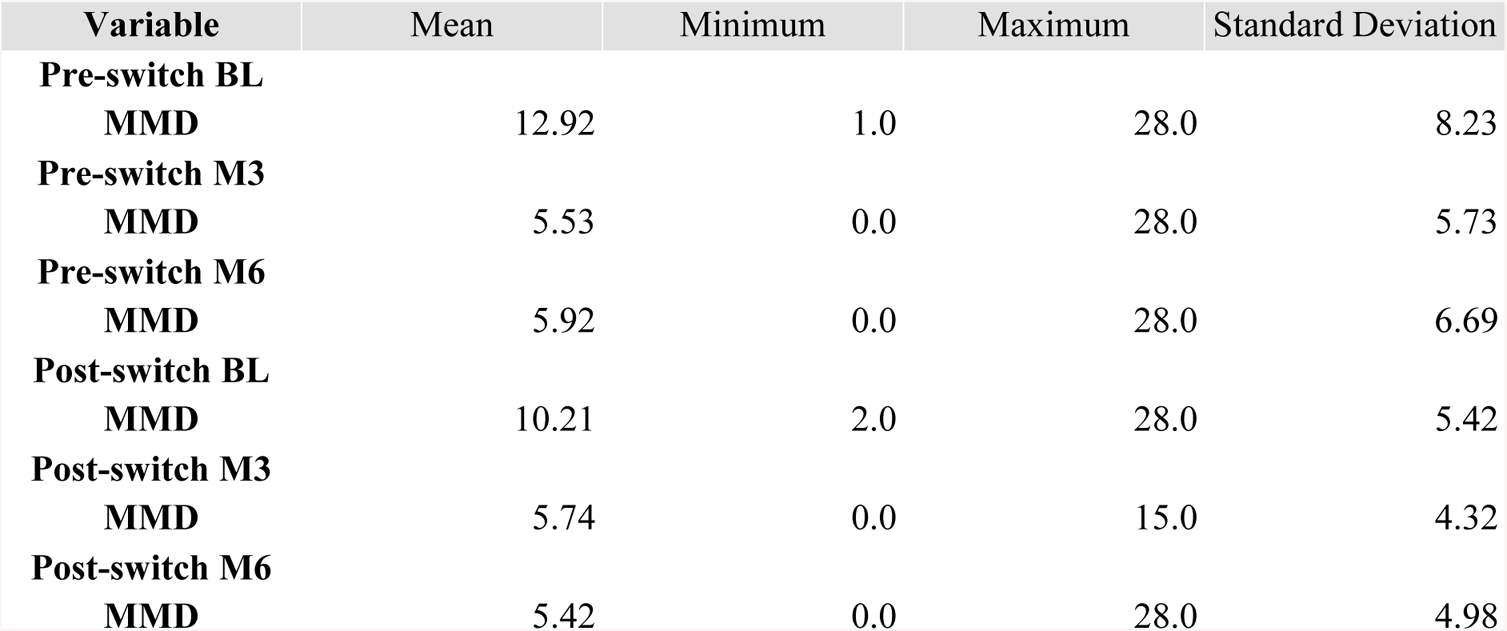
MMD Mean Data Pre- and Post-Switch.

### First anti-CGRP monoclonal antibody

A total of 29 patients were initiated on CGRP/L mAbs, while 24 patients were started on CGRP/R mAbs. Upon conducting a follow-up at month 3, it was observed that patients with CM exhibited a higher response rate as compared to patients with EM. Specifically, 72% of CM patients responded to treatment with CGRP/L mAb, whereas 100% of CM patients responded to CGRP/R mAb. On the other hand, 61% and 63% of EM patients responded to CGRP/L mAb and CGRP/R mAb respectively. However, it is worth noting that the CM group displayed higher rates of non-responsiveness when treated with CGRP/L mAb (27%) as opposed to CGRP/R mAb (0%). Upon analyzing the 6-month follow-up data, no significant improvement in response rates was observed. Notably, 8 EM patients remained non-responders to both classes of CGRP-mAb. Further details regarding response rates during the initial observational period on the first medication are displayed on **Fig 3**.

**Figure 3:**
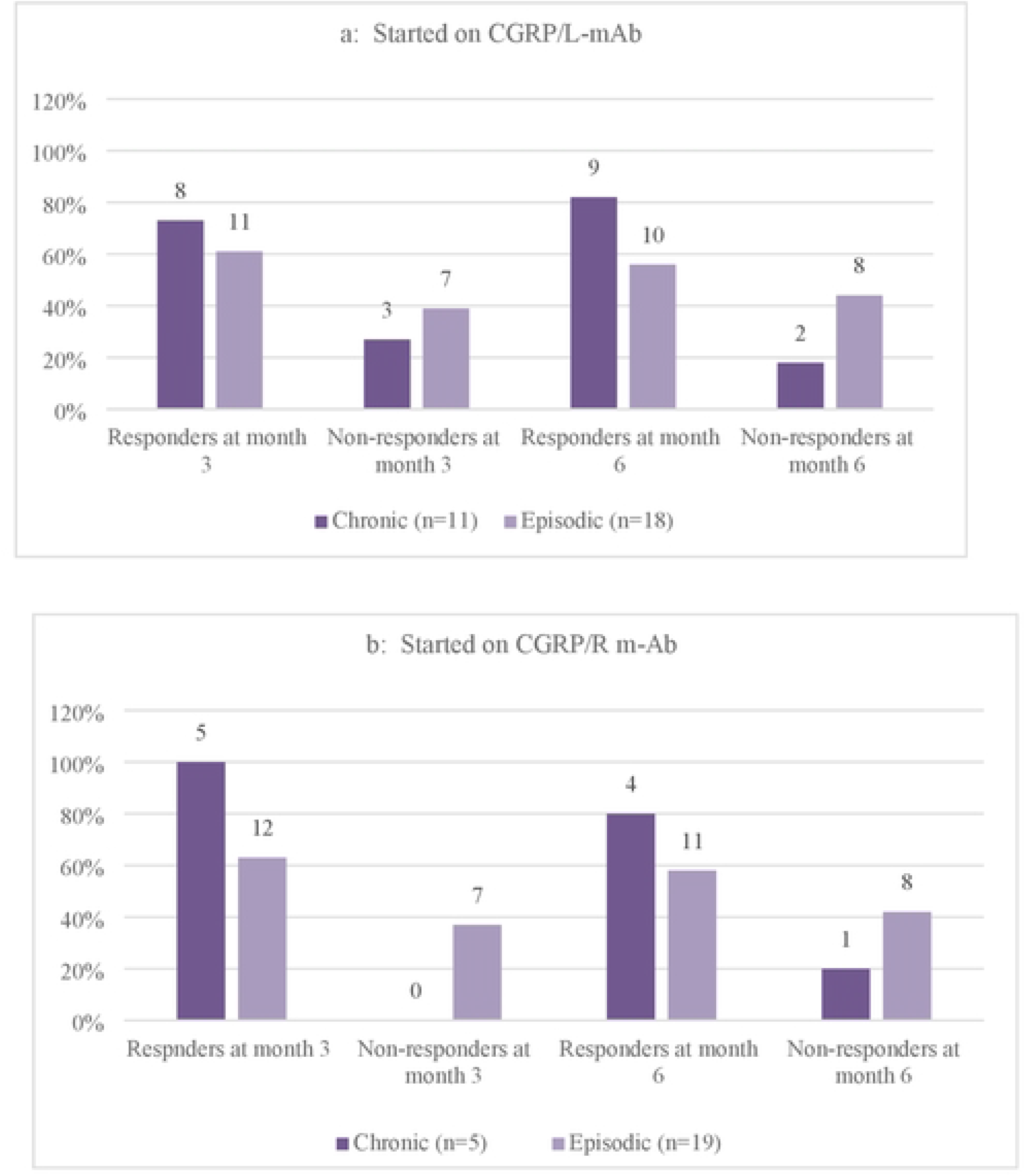
Responders and non-responders’ rates at month 3 and month 6 of patients with EM or CM on **3a:** CGRP/L mAb and **3b:** CGRP/R mAb dming the first observational period.

### Second anti-CGRP monoclonal antibody

During the second observational period, the 24 patients who received CGRP/R-mAb as their first CGRP-mAbs switched to anti-CGRP/L-mAbs (7 Galcanezumab and 17 Eptinezumab), whereas out of the 29 patients who started on CGRP/L-mAb (28 Galcanezumab and 1 Eptinezumab), 11 patients were switched to CGRP/R-mAbs (Erenumab) and 18 patients to another class of CGRP/L-mAb (galcanezumab). All the patients in the latter group have switched from Galcanezumab to Eptinezumab, while the one patient who initially received Eptinezumab was switched to Erenumab. Surprisingly, response rates during the second observational period at month 3 follow-up dropped in the CM and EM patients who were switched from CGRP/R-mAbs to CGRP/L-mAbs, from 100% and 63% to 71% and 41% respectively, (**Fig 3b and 4a**). On the other hand, CM patients who started on CGRP/L-mAb, had a higher response rate when they were switched to another anti-CGRP/L-mAb (100%) than to anti-CGRP/R-mAb (87.5%) at month 3, (**Fig 4b and 4c**). Overall, CM had a better response rate than EM during the second observational period **Fig 4**.

**Fig 4:**
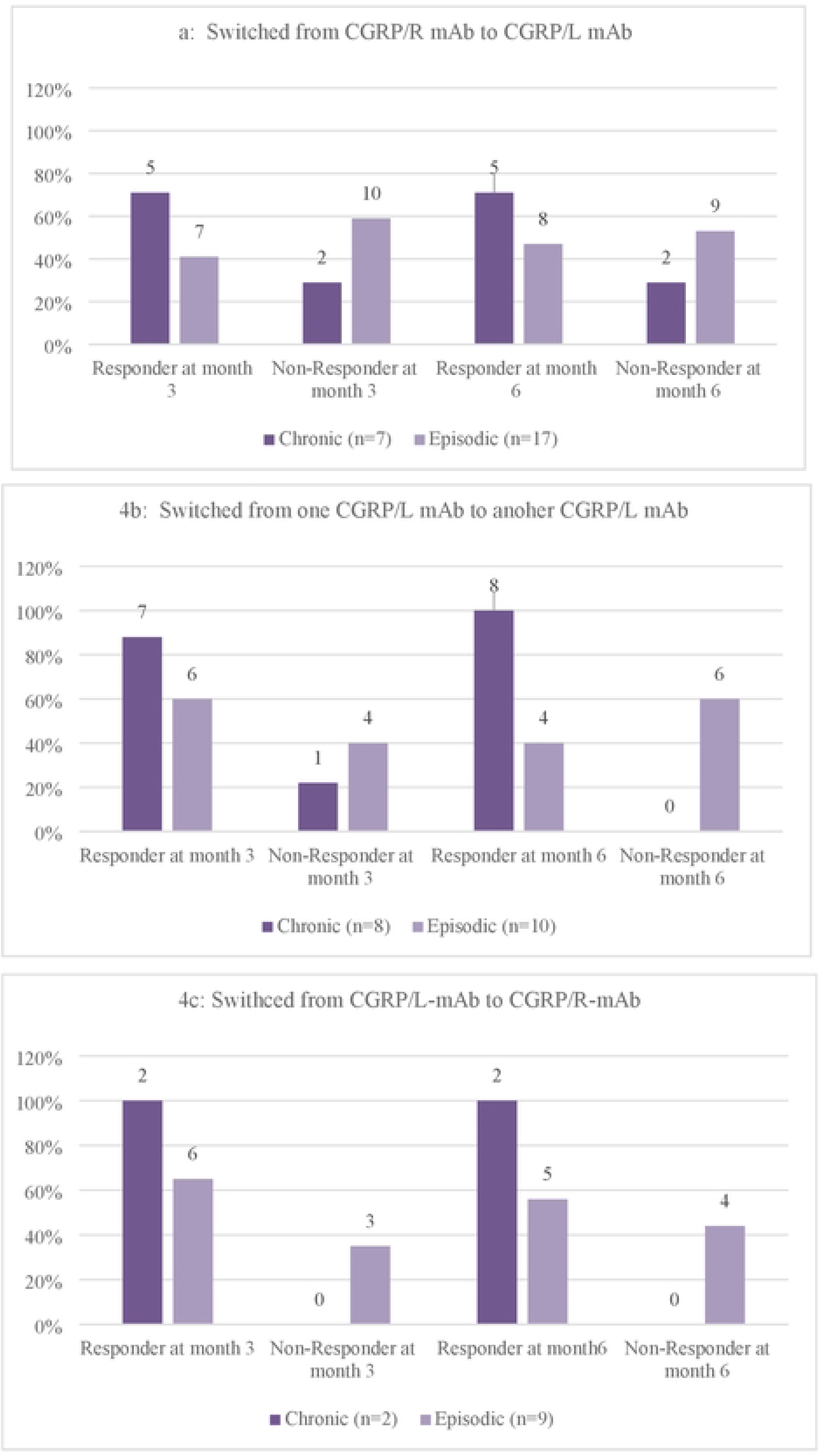
Responders and non-responders’ ratesat month 3 and month 6 of patients with EM or CM on **4a** after switching from CGRP/R-mAb to CGRP/L-mAb, **4b:** from CGRP/L-mAb to CGRP/L-mAb **4c:** Switched from CGRP/L-mAb to CGRP/R-mAb.

### First vs Second anti-CGRP monoclonal antibody

**Table 4** and **table 5** present the median differences in MMDs at 6 months compared to the baseline. These tables specifically focus on the first and second CGRP mAbs administered to different patient groups. Interestingly, the overall reduction in MMDs for all patients is precisely identical for both the first and second mAbs.

**Table 4:**
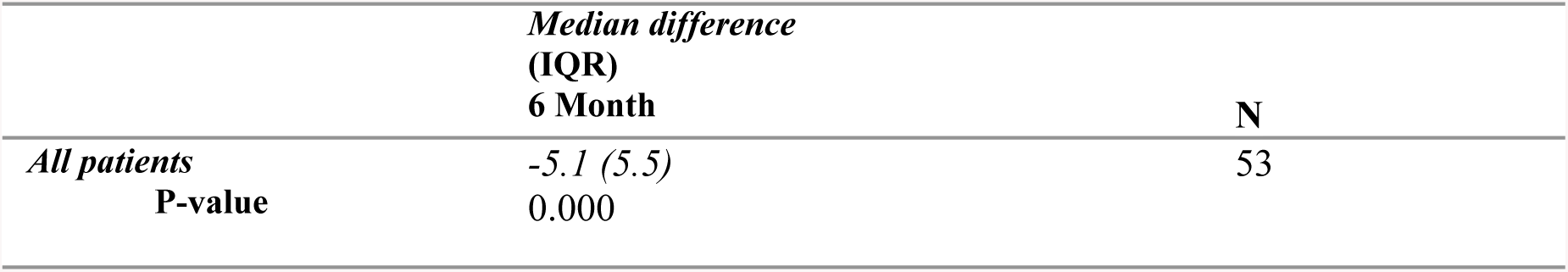
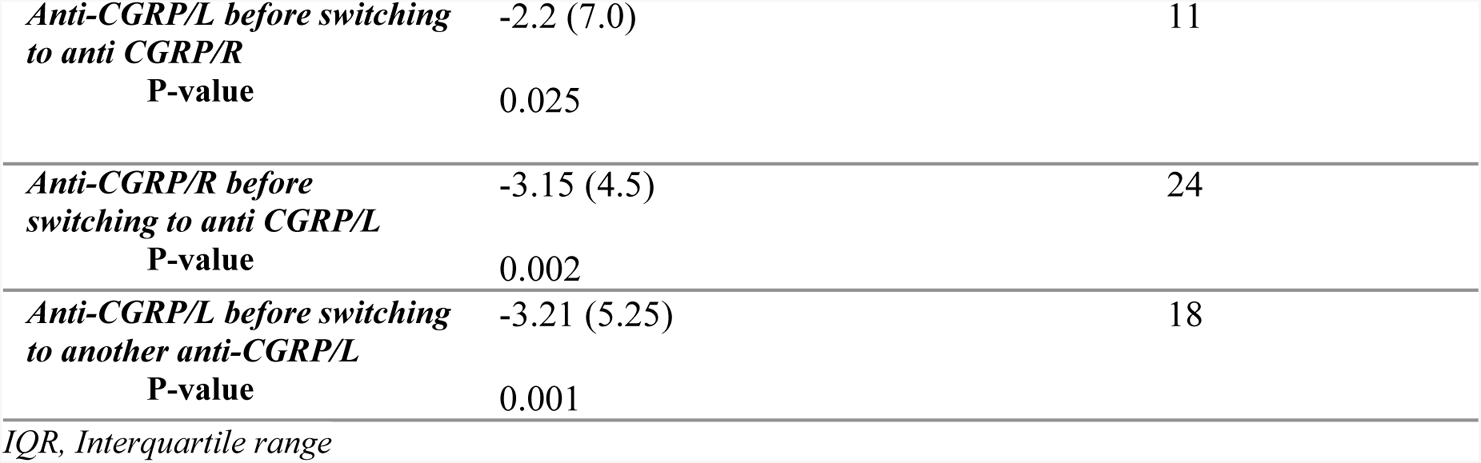
Changes in MMD during first CGRP mAb from 1^st^ baseline.

**Table 5:**
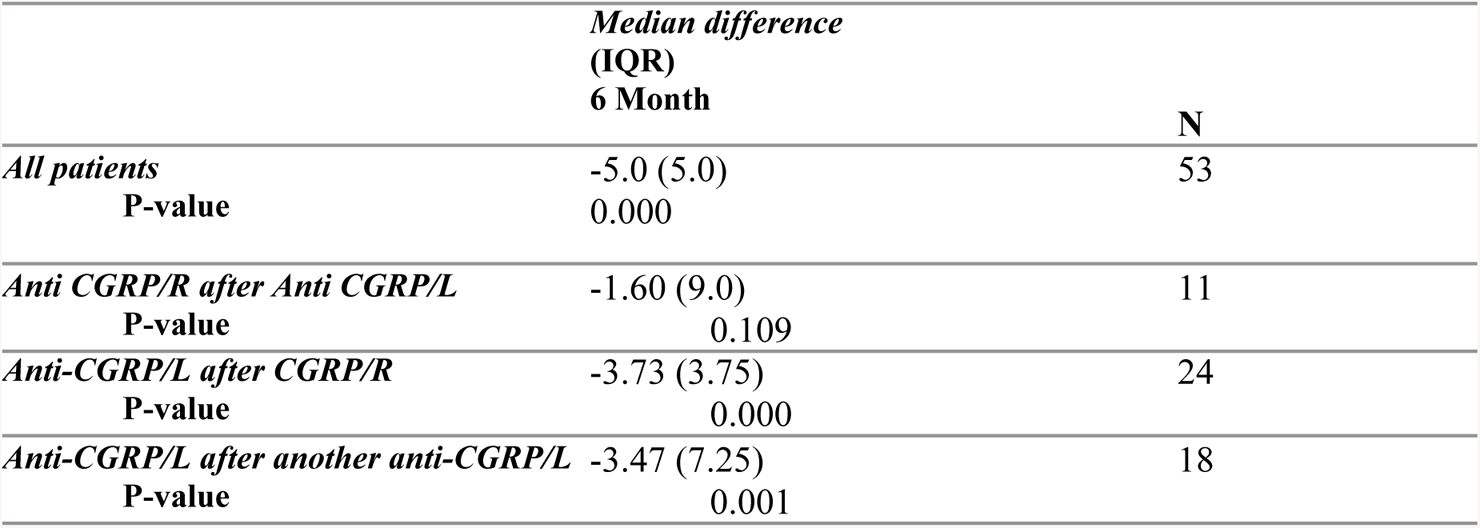
Changes in MMD during second CGRP mAb from 2^nd^ baseline.

Documented in **Table 6**, patients were assessed on their reduction in MMDs from the 6-month follow-up pre-switch to their 6-month follow-up post-switch as an additional evaluation of treatment efficacy following a medication switch. In line with the results of the Wilcoxon tests, among ligand-receptor switchers, only 1 (11.1%) participant experienced a greater than 50% reduction in MMDs, while the vast majority (66.7%) experienced less than 25% reduction. Although a greater proportion of receptor-ligand switchers (19.1%) experienced greater than 50% reduction on MMDs, an overwhelming majority (71.4%) experienced less than 25% reduction. Interestingly, the ligand-ligand cohort exhibited the greatest proportion (33.4%) of participants experiencing greater than 50% reduction, and the lowest proportion (44.4%) of those experiencing less than 25% reduction.

**Table 6:**
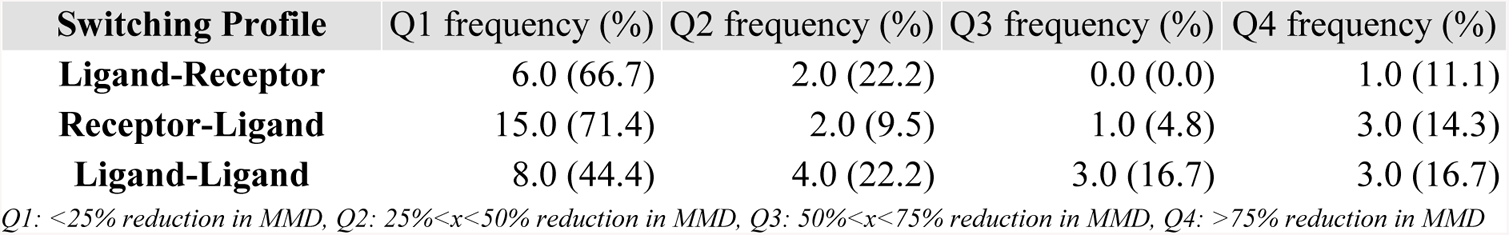
M6-M6 Percent Reductions.

### Exposure and Safety

In this particular study, a total of 53 patients were included. Out of these patients, it was observed that 4 individuals had an adverse event (AE), 3 of which took place prior to the switch in treatment. However, it is important to mention that despite these AEs, these patients did not opt to discontinue the treatment, indicating that the AEs were of a minor nature. AEs included constipation, slight pain on injection site, and increased itchiness. The remaining patient reported an AE after the switch. This particular AE involved peri labial numbness and swelling, which occurred during the infusion. The patient was closely monitored and subsequently discharged safely. Treatment was well-tolerated among the remaining 49 patients.

## Discussion

Despite the recent growth in CGRP-mAb use as a unique migraine treatment strategy, relatively little is known regarding how switching between CGRP-mAb classes can impact the efficacy and tolerability of treatment. Therefore, this study represents another step in furthering our understanding of how CGRP-mAb treatments can be safely and effectively applied in a clinical context. Moreover, to our knowledge, this is the first real-world study of its kind to be conducted in the United Arab Emirates and GCC region, including, this underrepresented population, from the pivotal trials. Determined prior to data analysis, our primary endpoint was to determine the impact, if any, of switching between CGRP-mAb medications on the effectiveness of MMD reduction. Furthermore, our secondary endpoint was to assess the safety and tolerability associated with that switching.

The 53 patients included in data analysis were classified according to their switching profile including: switching from receptor-targeted to ligand-targeted treatment (RL), from ligand-targeted to ligand-targeted (LL) treatment, and from ligand-targeted to receptor-targeted (LR) treatment. Using a Wilcoxon Signed Ranks Test, groups were compared at baseline (BL), month 3 (M3), and month 6 (M6) stages to assess if changes in MMDs were attributable to treatment. Although RL and LL patients experienced significant reductions in MMD between BL and M6 after switching, this was not the case for LR patients. This incongruence between groups warrants further investigation, with larger cohorts, into the pathophysiology of CGRP-mAb treatments to explain why switching in one direction produces starkly different outcomes than the other.

Indeed, prior to conducting the study, we expected switching between different classes of CGRP mAbs would yield improved results, especially for those switching from L to R or vice versa. However, to our surprise, our results showed that switching from ligand to ligand produced better outcomes. This unexpected finding underscores the importance of studying the mechanism of CGRP mAb treatments in greater detail. Conducting future studies with larger cohorts would provide a better insight into the pathophysiology of these treatments and may help further understand why switching from ligand to ligand produces a better outcome.

Despite a lack of statistically significant difference between M6 values pre- and post-switching, we found it valuable to quantify the percent MMD reduction according to switching profile. Notably, 33.4% of LL patients experienced at least 50% reduction in MMDs 6 months post-switch, when compared to their 6-month data from their initial medication. Among LR and RL patients, less than 20% of each group experienced a similar degree of MMD reduction. These findings offer modest support to the notion that considering drug mechanism of action may lend itself to improved outcomes post-switching. Unfortunately, as Erenumab is the only drug included targeting CGRP receptors, there is no means of assessing a receptor-receptor switch for a similar phenomenon.

On the topic of safety, 3 of 4 documented AEs took place prior to switching, all of which were considered mild and did not impact patients’ continuation on treatment. It is worth noting that despite experiencing AEs on their previous CGRP-mAb treatment, the aforementioned 3 patients switching to another mAb did not result in worsening or new AEs. This is a promising development pointing to switching being as a safe, tolerable, and, probably, effective process for those experiencing side effects with their first CGRP-mAb prescription.

We have identified two retrospective studies examining lateral switching between CGRP mAb therapies (20,21), to serve as points of comparison of our findings relative to those in the existing literature. The study by Overeem et al. (20) conducted a real-world, multicentre analysis of 78 patients with PPTF on erenumab (receptor-targeted therapy) who switched to ligand-targeted therapies. Unlike the 50 % meaningful response rate that was used in our study, their analysis yielded >30% reduction in MHDs by month 3 after switching in 32% of patients and >50% reduction in 12% of patients.

On the other hand, the study by Kaltseis et al. (20,21), conducted a larger retrospective assessment of 171 who received either one, two, or three different anti-CGRP mAbs. In contrast to the study by Overeem et al., non-response was set at <50% reduction in MMDS in EM patients, and <30% reduction in CM patients. (5.3%) of participants discontinued due to negative side effects. As compared to our study, that study was heavily focused on the quantity of PPTFs. Our study, however, has carefully analysed, the impact of switching directionality on outcomes.

In addition, and in contrast to the study by Overeem et al (20), where patients who switched from CGRP/R mAb to any CGRP/L mAb experienced improved MMDs, our study did not observe similar outcomes, which could be due to the different characteristics of our cohorts as compared to theirs. Indeed, our cohorts included 37 naïve patients who have not previously tried other preventive therapies, representing, probably, less refractory patients than those previously studied in the literature. Out of the 53 patients in our cohorts, 16 individuals had previously attempted preventative treatment for migraine.

### Limitations

Our study had several limitations that must be acknowledged. Being retrospective in nature means that controlling certain variables that may have affected the results was not possible. Additionally, the sample size included was small, which limits the generalizability of our findings. As a result, it is worth noting that the data analyzed involved one patient who was initially on Eptinezumab who have switched to Glacanezumab, and consequently further studies may be required to confirm our findings on Ligand-Ligand lateral switching outcomes following similar direction of switching, and to verify the generalizability of the data. Furthermore, the effectiveness outcome was solely based on MMD reduction. Thus, it may not fully capture the impact of the intervention on other patient related outcomes. Due to the relatively short observation period of six months for both first and second medication, it is not possible to rule out the potential for further improvement that could have been attained if patients had prolonged their use of the initial medication before switching to an alternative one. However, the recent European Headache Federation (EHF) guidelines have suggested 3 months of observation is sufficient to assess the effectiveness of mAbs (22). Finally, it is important to acknowledge the differences in regulation of prescribing mAbs in UAE as compared to other parts of the world. It is worth noting that currently in UAE there exists no prerequisite for individuals to have undergone a specific number of unsuccessful preventative treatment prior to being administered a CGRP mAb, which make our studied cohorts unique in that regards.

## Conclusion

This retrospective, real-world exploratory study examining the effectiveness and safety of switching between CGRP-mAb treatments serves as an essential step into furthering our understanding of an under-researched topic. The findings of this study suggest that the outcome of switching from a previous treatment does not significantly influence the outcome of future prescriptions’ effectiveness nor safety.

Among 53 patients enrolled, none experienced significant changes in their MMD when comparing their mean data 6 months on the previous medication against 6 months on the new medication. Nevertheless, the data suggests that those switching from one ligand-targeted treatment to another ligand-targeted treatment were more likely to experience additional or compounded reductions in MMD on top of improvements gained from their initial prescription.

Overall, the findings present in this study point to CGRP-switching as a potentially safe and clinically viable practice that may have applications for those experiencing suboptimal response on their current CGRP-mAb. Further research is warranted to better understand the long-term implications of switching beyond a 6-month period, as well as if those switching to a CGRP-mAb of the same class are truly likely to experience greater improvements than their counterparts.

## Data Availability

All relevant data are within the manuscript and its Supporting Information files.

